# Comparing Sulfadoxine-Pyrimethamine+Chloroquine and Dihydroartemisinin-Piperaquine to Control for Malaria Prevention in Malawian School Children: Results from a Randomized Controlled Trial

**DOI:** 10.64898/2026.08.04.26359752

**Authors:** Wongani Nyangulu, Enalla Mzembe, Wangisani Kumalakwaanthu, Alfred Matengeni, Alick Sixpence, James Chirombo, Miriam K. Laufer, Don P. Mathanga, Lauren M. Cohee

## Abstract

Malaria remains a significant global health challenge. Intermittent Preventive Treatment of malaria in school-age children (IPTsc) is recommended to reduce disease burden, but optimal drug choice is unclear. Dihydroartemisinin-Piperaquine (DP) is highly efficacious but there are concerns about widespread use given emerging artemisinin resistance and its role as an alternative first line treatment. Thus, non-artemisinin alternatives to DP are needed.

646 Malawian primary school children participated in the second stage of a 3-arm randomized controlled trial. Participants were allocated to IPTsc with 1) DP, 2) Sulfadoxine-Pyrimethamine+Chloroquine (SP+CQ) or 3) Control (no treatment). Study drugs were administered at three six-weekly visits. Outcomes were measured 6–8 weeks later. The primary outcome was *Plasmodium falciparum* (*Pf*) prevalence detected by qPCR. Secondary outcomes included clinical malaria and anemia. Analysis was modified intention-to-treat.

Outcome assessment included 588 (91%) participants. Prevalence of *Pf* infection was 18% (34/198) in the IPTsc-DP arm, 27% (52/200) in the IPTsc-SP+CQ arm, and 48% (89/190) in the control arm. Compared to control, both IPTsc-DP (adjusted Odds Ratio [aOR] 0.22, 95%CI:0.14–0.36, p<0.001) and IPT-SP+CQ (aOR 0.38, 95%CI:0.24–0.59, p<0.001) significantly reduced odds of infection. Both regimens also decreased anemia (DP: aOR 0.45, 95%CI:0.21–0.93, p=0.035; SP+CQ: aOR 0.47, 95%CI:0.22–0.98, p=0.048) and clinical malaria (DP: adjusted Incidence Rate Ratio [aIRR] 0.41, 95%CI:0.28–0.60), p<0.001; SP+CQ: aIRR 0.60, 95%CI:0.43 – 0.84, p=0.003).

In Malawi and settings with similar malaria drug resistance profiles, SP+CQ may be a suitable alternative to DP for IPTsc.

**Clinical Trial Registration:** ClinicalTrials.gov ID: NCT05980156

## Introduction

Malaria remains a major global health challenge causing an estimated 282 million cases and 610,000 in 2024 [1]. The vast majority of this burden (95% of cases and deaths) falls on African countries. Primary school age children, typically 5–15 years old, bear a significant yet underappreciated burden of malaria. In moderate to high transmission settings, they have the highest prevalence of infection compared to both younger children and adults [2–4]. Most infections in this age group present as either acute uncomplicated clinical malaria episodes or chronic sub-clinical infections which are associated with anaemia, poor cognitive function, and reduced school attendance [2, 5–7].

Intermittent Preventive Treatment (IPT), which is a form of malaria chemoprevention, utilizes a full course of anti-malarial drugs to clear current infections and provide a period of post-treatment prophylaxis, the duration of which depends on drug half-life. IPT has been successfully used in pregnant women and young children [8, 9], and clinical trials have demonstrated that it prevents infection, clinical malaria episodes, anemia and cognitive impairment in school children [10, 11]. While the World Health Organization issued a conditional recommendation for IPT in school-age children (IPTsc) [12], adoption has been limited due to unresolved questions, including optimal drug choice in the context of existing and emerging drug resistance. Dihydroartemisinin-piperaquine (DP), a long-acting artemisinin-combination therapy (ACT), has emerged as the most effective drug for IPTsc [13, 14]. While current guidelines do not specify which drugs should be used for IPTsc, they state that first-line treatment drugs should not be used for chemoprevention.

Drug resistance has shaped the choice of malaria treatment and prevention drugs for the last three decades. In 1993, Malawi was the first country to remove chloroquine as first-line treatment for malaria on the basis of extensive drug resistance, replacing it with sulfadoxine-pyrimethamine (SP) [15–17]. More than a decade later, in 2007, Malawi adopted artemisinin-combination therapies (ACTs) for first-line treatment due to the development of SP resistance [18, 19]. Artemisinin resistance, which first emerged in Southeast Asia [20], has now been detected in several East African countries [21, 22]. While ACTs remain clinically efficacious even where there are markers of resistance due to the absence of resistance to partner drugs, reports in the Democratic Republic of Congo, Angola, Burkina Faso and Uganda raise concerns [22]. In Malawi, growing concerns over the declining efficacy of artemether lumefantrine, the ACT currently used as first-line treatment for uncomplicated malaria, have prompted the adoption of DP as an alternative first-line regimen [23–25]. Thus, alternative non-artemisinin options for chemoprevention, including IPTsc, are needed.

Following its removal from the market, susceptibility to chloroquine (CQ) has returned in Malawi [15]. In contrast high prevalence of molecular markers of SP-resistance persist [26, 27]. However, the antimalarial efficacy of SP in semi-immune populations without clinical illness is not clear [28]. We aimed to evaluate SP+CQ as a potential, non-artemisinin drug regimen for IPTsc. We extended a trial which demonstrated that IPTsc was superior to Intermittent Screening and Treatment (IST) [29] and hypothesized that both SP+CQ and DP would be effective drug regimens to decrease malaria infections, clinical malaria disease, and anaemia compared to control. This comparison will inform policy on malaria prevention targeting school-age children, which has the potential to improve health and education outcomes in children and advance malaria elimination efforts.

## Methods

### Study design and setting

From February to July 2023, students in Nainunje Primary School in Machinga district, Malawi, participated in the continuation of a three-arm, individually randomized, open-label clinical trial (clinicaltrials.gov: NCT05980156). The study protocol was reviewed and approved by the Kamuzu University of Health Sciences Research Ethics Committee (P.06/21/3410) and the University of Maryland School of Medicine (HCR-HP-00098250-2). Briefly, the study area is rural with high malaria transmission, including a seasonal peak between January and May. Site selection criteria and results from the initial trial were previously published [29].

### Participants

All participants from the initial trial were offered enrolment in the continuation. Written informed consent and assent (from participants who were 10 years or older) were obtained before participation. Exclusion criteria included: current evidence of severe malaria or danger signs, known adverse reaction to study drugs, history of cardiac problems or fainting, current treatment with medications known to prolong QT interval, family history of prolonged QT interval or unexplained sudden death, epilepsy, or psoriasis.

### Randomization and masking

Participants were randomized in the initial study and remained in the same group in the continuation, though the treatment assignment changed for the IST arm. The trial was open-label, but laboratory technicians conducting parasite detection were blinded to the study allocation.

### Study procedures and data collection

In the continuation, participants in the IPTsc-DP arm in the initial study again received IPT with DP (or CQ for girls ≥ 10 years old). Participants in the intermittent screening and treatment arm in the initial study received IPT with SP+CQ (or CQ only for girls ≥ 10 years old) in the continuation phase. Participants in the control arm in the initial study continued in the control arm. At the end of the initial study (∼seven months before the start of the continuation phase), participants in the control arm were screened once for infection using a malaria rapid diagnostic test (NxTek™ ELIMINATE MALARIA Pf, Abbot Diagnostics Korea, Inc.) and treated if positive. Older girls (≥ 10 years old) who were likely to have started menses were treated with CQ only due to safety concerns about the use of DP and SP in the first trimester of pregnancy.

Following enrolment in the continuation phase, caregivers were interviewed to assess household-level insecticide-treated net ownership, education level of the household head, and socioeconomic status (SES) using ownership of assets, livestock, type of fuel used in the home and food security, which we report as an inverse frequency-weighted wealth index divided into tertiles [30].

Following enrolment, three intervention visits were conducted at six-weekly intervals during the peak malaria transmission season, and an outcome visit was conducted 6-8 weeks after the last intervention visit. At each intervention visit, participants allocated to the treatment arms received study drugs administered using weight-based dosing (Supplemental Table S1) as follows: Dihydroartemisinin-piperaquine (DP, P-ALAXIN, BLISS GVS PHARMA LTD, India) daily for three days, Sulfadoxine-pyrimethamine (SP) single dose, and Hydroxychloroquine (CQ, Zentiva Pharma UK Limited, United Kingdom) full dose on days one and two, half dose on day three. At each visit, the study team interviewed participants to collect data on bed net use, well-being, and fever in the last 48 hours. At the first intervention visit and the outcome visit, staff also collected finger-prick blood samples. Two 50 μL samples were placed on Whatmann 3MM filter paper, and another sample was used to measure hemoglobin (HemoCue 301). All data were collected on tablet computers. Adverse events were monitored by the study team following each visit and passively at the local health centre. Safety data were reviewed by a local monitor.

### Outcomes

The primary outcome was the detection of *Plasmodium falciparum* by PCR. Briefly, DNA was extracted from filter papers and *Pf* ribosomal RNA 18S gene was detected and quantified by qPCR[31, 32]. Secondary outcomes were anemia, hemoglobin and incidence of clinical malaria. Anemia was defined using the WHO age and sex specific cut-offs: for children <12 years old, hemoglobin <11.5 g/dl; for all children 12–14 and females 15 and older, hemoglobin <12.0 g/dl; and for males 15 and older, hemoglobin <13.0 g/dl [33]. Clinical malaria was defined as seeking clinical care, having a positive malaria RDT, being prescribed antimalarial treatment by a healthcare worker, and occurring more than 28 days after a prior clinical diagnosis. Cases that occurred between the first intervention visit and outcome visit were captured by passive case detection at the local health center and review of the participants’ health passport (portable medical record) to ensure all clinical malaria diagnoses were documented.

### Statistical methods

The sample size (n=750) was determined for the initial trial to have 80% power to detect a 40% relative reduction in infection prevalence [29]. Data were analysed using modified intention-to-treat (mITT), where all participants who had outcome measures available were included in the analysis, and only those participants who did not attend the outcome visit were excluded. Baseline characteristics were compared across the arms because baseline values of outcome measures might differ based on interventions received in the initial phase of the study. Categorical baseline characteristics were summarized using proportions and compared using chi-squared tests or Fisher’s exact tests, followed by pairwise comparisons.

Continuous baseline characteristics were summarized using means and compared using analysis of variance (ANOVA) with post hoc Tukey test and Scheffe tests done for pairwise comparisons. We adjusted for multiple comparisons using the Bonferroni correction.

For the primary outcome, presence of *Pf* infection and the secondary outcome, anemia, each intervention arm was compared to the control arm using univariate and multivariable logistic regression analysis. For the primary outcome model (*Pf* infection), we adjusted for grade level alone while for the secondary outcome (anemia), we adjusted for grade level and baseline anemia. Hemoglobin levels at the outcome were compared using an ANCOVA model where hemoglobin at the outcome visit was the outcome variable, study arm was the categorical predictor, grade level, and baseline hemoglobin were the covariates. The incidence of clinical malaria was compared using a Poisson regression model with log link offsetting for the log of the number of follow-up days and adjusting for grade level to obtain incident rate ratios (IRR). We calculated the treatment efficacy for preventing clinical malaria and the number needed to treat to prevent a single clinical malaria episode. We then compared clinical malaria-free survival outcomes across the study arms using Kaplan-Meier survival curves with log-rank tests. To adjust for potential confounding, we further assessed clinical malaria-free survival using a Cox proportional hazards regression model. We adjusted for grade level and baseline *Pf* infection.

## Results

Among the 746 children in the initial study, 646 (87%) enrolled in the continuation phase. Reasons for not enrolling included 78 students who graduated from primary school, 19 who withdrew consent, transferred out of the study school or were lost to follow-up during the initial study, and three who did not consent to participate in the continuation (Figure 1). More than half (54%, 352/646) of enrolled participants were female, and the mean age was 12.04 years (SD 2.97) (Table 1). At baseline, the overall prevalence of *P. falciparum* detected by PCR was 53% and the prevalence of anaemia was 23%. In pairwise comparisons, baseline *Pf* prevalence was borderline higher for IPTsc-DP arm compared to IPTsc-SP+CQ arm, but there were no significant differences between the other variables (Supplementary Table S2).

**Figure 1:**
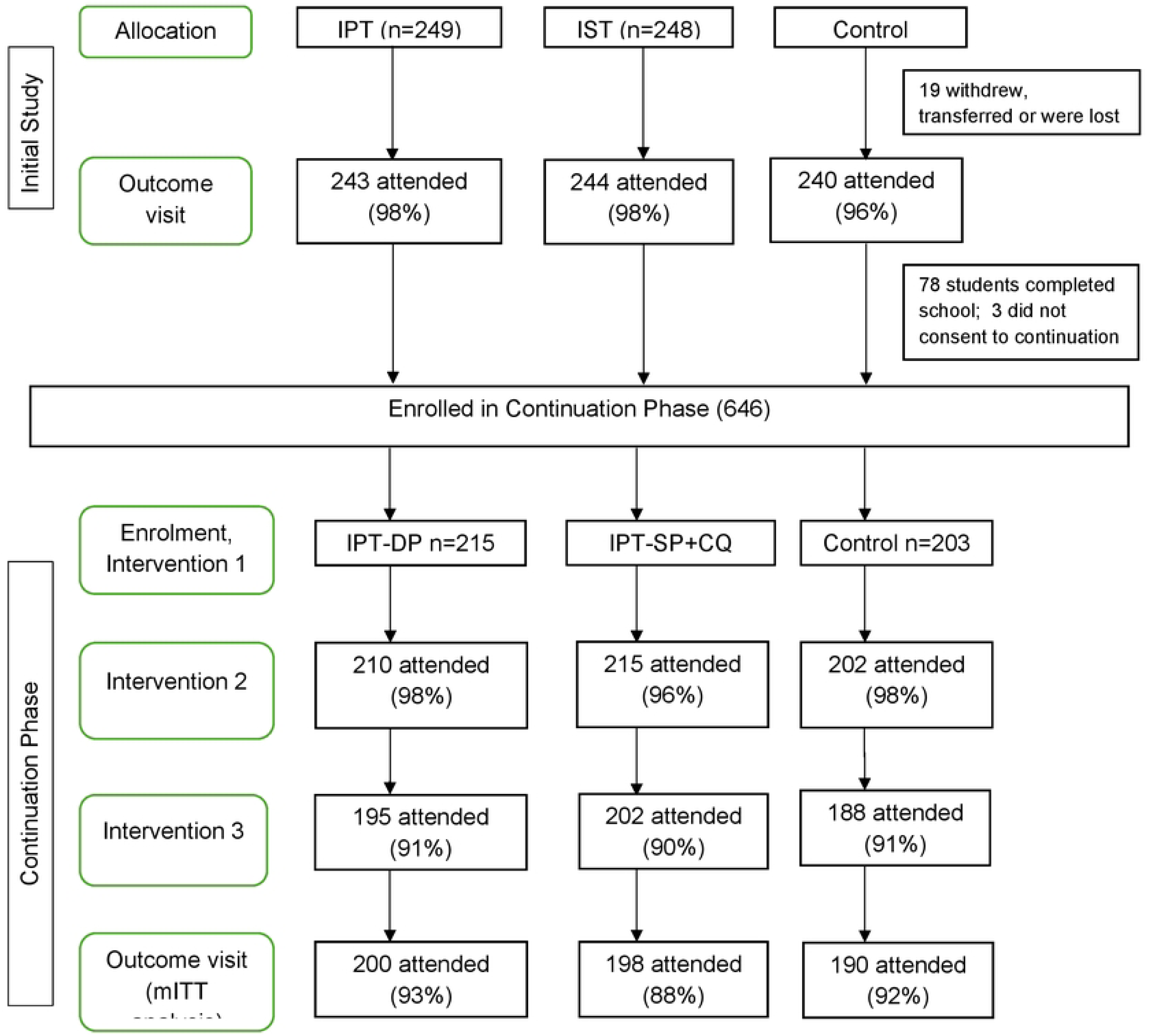
Trial profile. IPT Intermittent Preventive Treatment; 1ST Intermittent Screening and Treatment; DP Dihydroartemisinin-Piperaquine; SP Sulfadoxine-Pyrimethamine; CQ Chloroquine; mITT modified Intention to Treat. Following the first phase, 646 children consented to participate in the continuation phase. 588 (91%) participants attended the outcome visit at the end of the continuation phase, while 58 did not. Reasons for not attending intervention visits in the continuation phase included absent for unknown reasons (n = 421), withdrew consent (n = 17), already on malaria treatment (n = 15), absent due to illness (1), travel (2), refused for religious reasons (1) and others (90). Among these, 36 said it was the end of the week, and they could not come for the visit, 17 transferred to another school and 11 relocated to find work on farms, to Mozambique or were displaced by Cyclone Freddy, 10 missed treatment, 7 became pregnant, 6 stopped school, 2 were on malaria treatment and 1 was very ill and could not attend the visit.

**Table 1.**
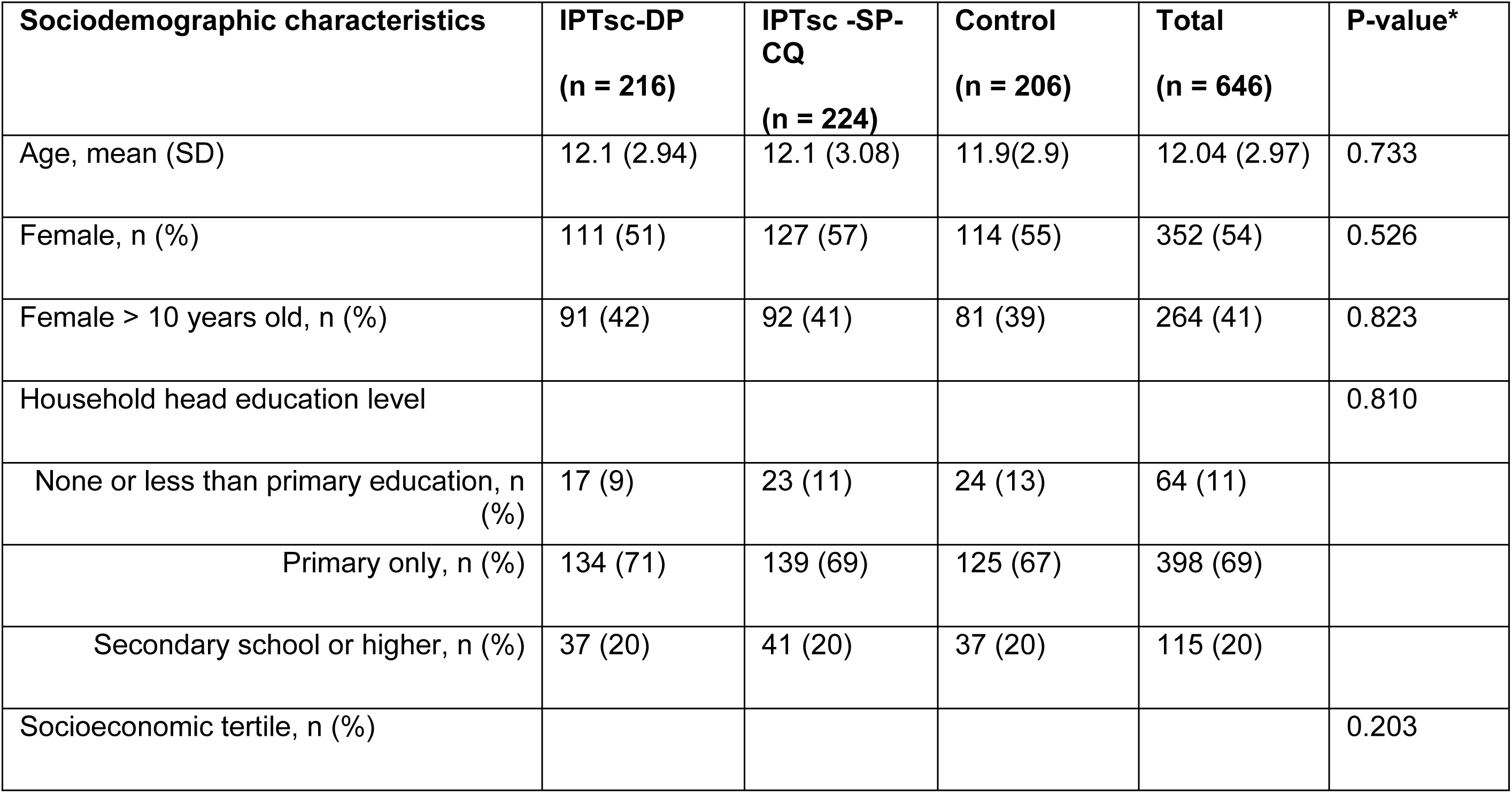
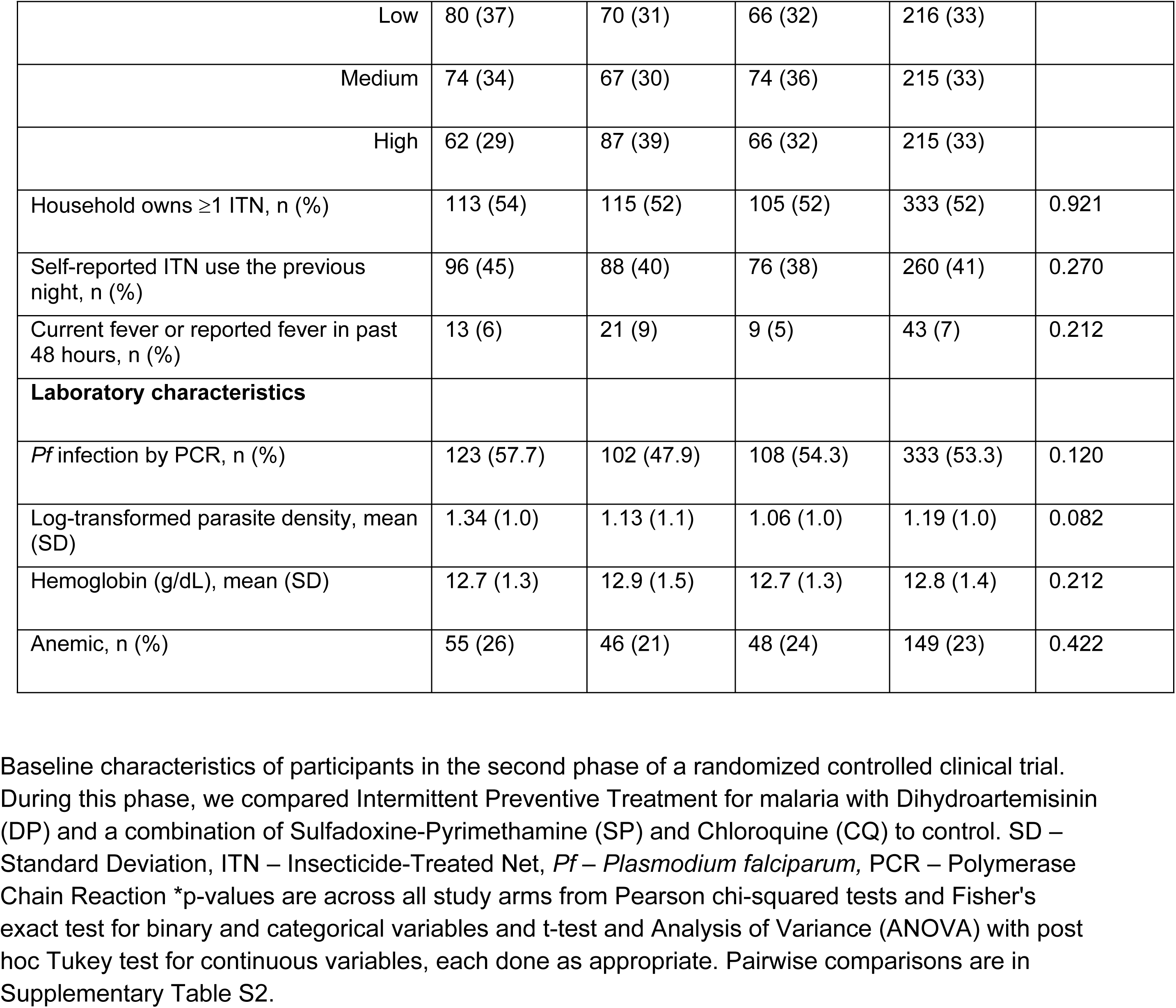
Baseline characteristics by allocated study arm of students participating in the continuation of a randomized clinical trial to evaluate drug regimens for intermittent preventive treatment in school-age children (IPTsc).

Participation in the intervention visits was high, with 96% of all possible visits attended (1854 attended visits/1938 possible visits) across the study arms. Among the attended intervention visits, 82% (1523/1854) were complete visits with all three days of study drugs administered. Treatment compliance was higher in the IPT-DP arm compared to the IPTsc-SP+CQ arm: 82% (495/607) of those who received treatment completed all three doses in the IPTsc-DP arm compared to 70% (438/624) in the IPTsc-SP+CQ arm (p<0.001; Supplementary Table S3).

Ninety-one per cent (588/646) of participants were evaluated at the outcome assessment and, thus, were included in the mITT analysis. The participants who did not attend the outcome visit (58 (9%)) were older and more likely to be male compared to those who attended the outcome visit, but there were no differences in the proportion of females >10 years old, socioeconomic status, household head education level, or bed net ownership (Supplementary Table S4).

At the outcome visit, prevalence of *Pf* infection was 18% (34/198) in the IPTsc-DP arm, 27% (52/200) in the IPTsc-SP+CQ arm, and 48% (89/190) in the control arm (Table 2). Compared to control, both IPTsc-DP (adjusted Odds Ratio [aOR] 0.22, 95%CI:0.14–0.36, p<0.001) and IPTsc-SP+CQ (aOR 0.38, 95%CI:0.24 – 0.59, p<0.001) significantly reduced the odds of infection. Among participants with infections detected at the outcome visit, there were no differences in the parasite density by study arm. The incidence rate of clinical malaria was 0.48 cases per person-year (95%CI:0.32 – 0.64) in the IPTsc-DP arm, 0.70 cases per person-year (95%CI: 0.51 – 0.89) in the IPTsc-SP+CQ arm and 1.17 cases per person-year (95%CI: 0.90 – 1.44) in the control arm. Both IPTsc-DP (aIRR 0.41, 95%CI:0.28 – 0.60, p<0.001) and IPTsc-SP+CQ (aIRR 0.60, 95%CI:0.43 – 0.84, p=0.003) significantly reduced the incidence of clinical malaria compared to control. The efficacy for preventing clinical malaria episodes was 58.8% (95%CI:39.9 – 72.3) in the IPTsc-DP arm and 40.1% (95%CI:16.3 – 57.4) in the IPTsc-SP+CQ arm. The number needed to treat to prevent one clinical malaria episode was 1.45 for IPTsc-DP and 2.13 for IPTsc-SP+CQ (Supplementary Table S5).

**Table 2.** The effect of three rounds of intermittent preventive treatment of school-age children with dihydroartemisinin-piperaquine (IPTsc-DP) and sulfadoxine-pyrimethamine + chloroquine (IPTsc-SP+CQ) on health outcomes

| Outcome | Control<br>N= 190 |  | IPTsc-DP<br>N = 198 |  |  |  | IPTsc-SP+CQ<br>N = 200 |  |  |  | IPTsc-DP vs IPTsc-SP+CQ |  |
| --- | --- | --- | --- | --- | --- | --- | --- | --- | --- | --- | --- | --- |
|  | N | (%) | N | (%) | aOR (95%<br>CI) | P-value | N | (%) | aOR (95%<br>CI) | P-value | aOR (95%<br>CI) | P-value |
| *Pf* by PCR | 89 | 48.4 | 34 | 18.1 | 0.22 (0.14<br>– 0.36) | <0.001 | 52 | 27.1 | 0.38 (0.24<br>– 0.59) | < 0.001 | 0.53 (0.32<br>– 0.88) | 0.020 |
| Anaemia | 25 | 13.2 | 14 | 7 | 0.45 (0.21<br>– 0.93) | 0.035 | 14 | 7.1 | 0.47 (0.22<br>– 0.98) | 0.048 | 0.95 (0.42<br>– 2.16) | 0.91 |
|  | Mean | SD | Mean | SD | Mean<br>difference<br>vs control<br>(95% CI) | P-value | Mean | SD | Mean<br>difference<br>vs control<br>(95% CI) | P-value | Mean<br>difference<br>(95% CI) | P-value |
| Hemoglobin<br>(g/dL) | 13.2 | 1.29 | 13.6 | 1.32 | 0.40 (0.14<br>– 0.67) | 0.001 | 13.6 | 1.37 | 0.34 (0.07<br>– 0.61) | 0.007 | 0.06 (-0.32<br>– 0.20) | 0.852 |
| Parasite<br>density (log-transformed) | 1.21 | 0.90 | 1.24 | 0.93 | 0.03 (-0.44<br>– 0.51) | 0.99 | 1.12 | 1.17 | -0.09 (-<br>0.50 –<br>0.32) | 0.86 | -0.12 (-<br>0.64 –<br>0.40) | 0.84 |
|  | Cases | Incidence<br>Rate (95%<br>CI) * | Cases | Incidence<br>Rate (95%<br>CI) * | aIRR (95%<br>CI) | P-value | Cases | Incidence<br>Rate (95%<br>CI) * | aIRR (95%<br>CI) | P-value | aIRR (95%<br>CI) | P-value |
| Clinical<br>malaria | 86 | 1.17 (0.90<br>– 1.44) | 37 | 0.48 (0.32<br>– 0.64) | 0.41 (0.28<br>– 0.60) | < 0.001 | 56 | 0.70 (0.51<br>– 0.89) | 0.60 (0.43<br>– 0.84) | 0.003 | 0.69 (0.45<br>– 1.04) | 0.070 |

SD: Standard Deviation; *Pf. Plasmodium falciparum*; PCR: Polymerase Chain Reaction; (\*) Adjusted incidence rates per person-year
For *Pf* detected by PCR and anaemia, univariate and multivariable logistic regression were done to compare each arm against the control. In the multivariable models, we adjusted for grade level alone (*Pf* by PCR) and grade level and baseline anaemia (anaemia). Parasite density was compared across the study arms using ANOVA with parasite density as the outcome variable and study arm as the categorical predictor. Post hoc tests were done using Tukey and Scheffe tests. Haemoglobin levels at the outcome were compared using an ANCOVA model where haemoglobin at the outcome visit was the outcome variable, study arm was the categorical predictor, grade level, and baseline haemoglobin were the covariates. The incidence of clinical malaria was compared using a Poisson regression model with log link offsetting for the log of the number of follow-up days and adjusting for grade level.

The Kaplan-Meier survival curve for clinical malaria disease demonstrated significant differences in malaria-free survival across the study arms (log-rank test, p < 0.0001) (Figure 2). Participants in the IPTsc-DP group experienced longer malaria-free survival compared to both the IPTsc-SP+CQ and control groups. Participants in the IPTsc-SP+CQ arm experienced longer malaria-free survival compared to those in the control arm. In the Cox proportional hazards regression model, both IPTsc-DP (aHR 0.39, 95%CI:0.26 – 0.61, p <0.001) and IPTsc-SP+CQ (aHR 0.57, 95%CI:0.39 – 0.84, p = 0.004) significantly reduced the hazard of malaria compared to control.

**Figure 2.**
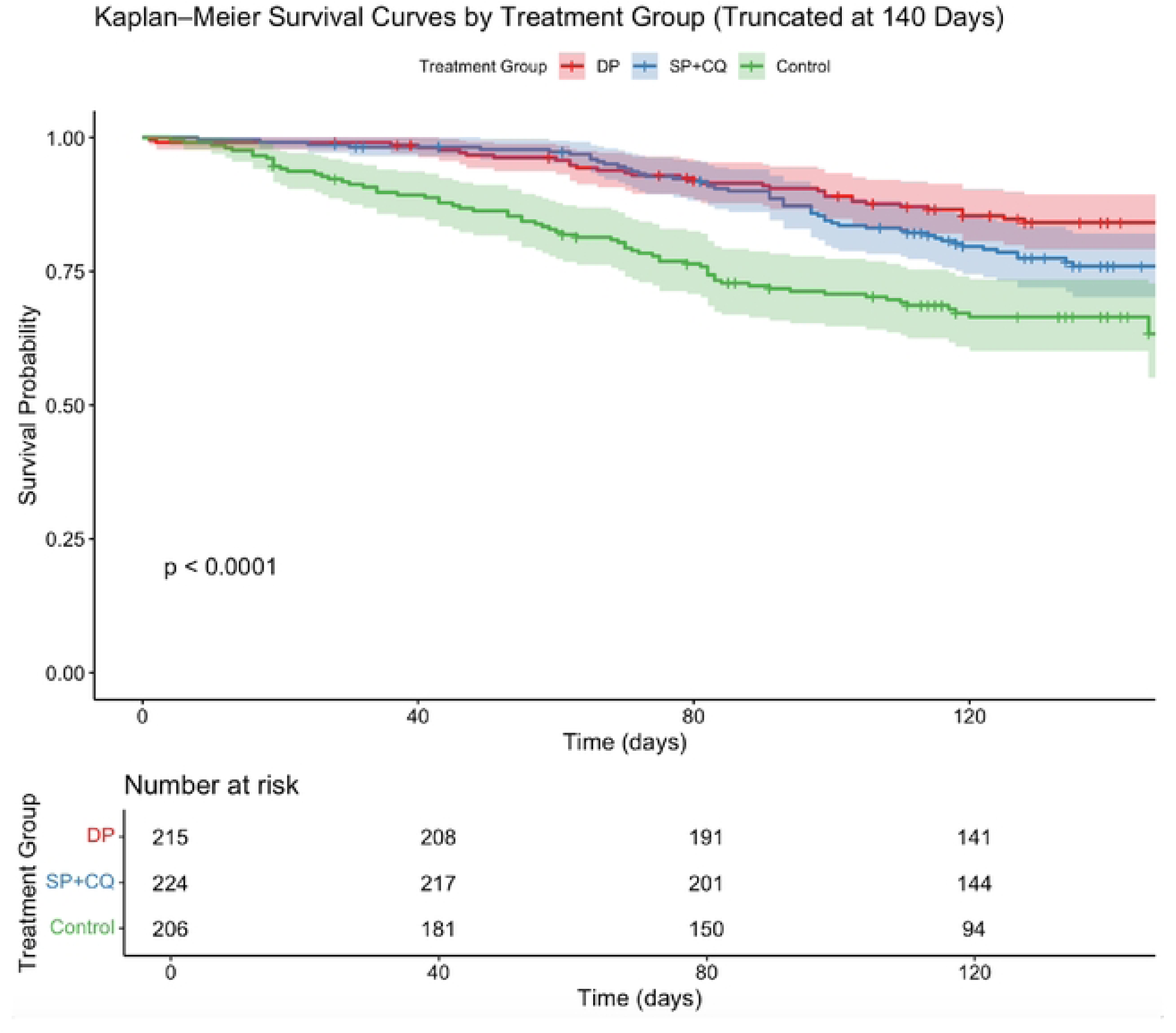
Kaplan-Meier survival curve comparing time to clinical malaria episodes between the study arms.

The mean hemoglobin concentration at the outcome was 13.6g/dL in the IPTsc-DP arm, 13.6g/dL in the IPTsc-SP+CQ arm, and 13.2g/dL in the control arm. Compared to control, both IPTsc-DP arm (adjusted mean difference 0.40g/dL, p=0.001) and IPTsc-SP+CQ (adjusted mean difference 0.34g/dL, p=0.007) significantly increased hemoglobin concentration compared to the control arm. Prevalence of anaemia was 7% (14/198) in the IPTsc-DP arm, 7% (14/200) in the IPTsc-SP+CQ arm, and 13% (25/190) in the control arm. Compared to control, both IPTsc-DP (aOR 0.45, 95%CI:0.21 – 0.93, p = 0.035) and IPTsc-SP+CQ (aOR 0.47, 95%CI:0.22 – 0.98, p = 0.048) reduced the odds of anemia.

While the trial was not designed to compare outcomes between the two intervention arms, IPTsc-DP significantly reduced *Pf* prevalence compared to IPTsc-SP+CQ (aOR 0.53, 95%CI:0.32 – 0.88, p=0.020). IPTsc-DP also showed a borderline reduction in incidence of clinical malaria compared to IPTsc-SP+CQ (aIRR 0.69, 95%CI:0.45 – 1.04, p=0.070). There was no difference between the groups in terms of the mean difference in hemoglobin and prevalence of anemia.

There were no serious adverse events reported in the study. There were sixty-five grade 1 and 2 (mild to moderate) adverse events reported by 3.2% of the participants (21/646) (Table 3). Most adverse events occurred in the IPTsc-SP+CQ arm and among participants who received chloroquine. The most common adverse events were abdominal pain, dizziness, fever, headache and nausea.

**Table 3.** Frequency of adverse events by study arm and drugs received as reported by participants who experienced them

| Symptom | Total, n (%) | By study arm, n (%) |  |  | By drug received, n (%) |  |  |
| --- | --- | --- | --- | --- | --- | --- | --- |
|  |  | IPT-DP | IPT-SP+CQ | IPT-Control | DP | SP+CQ | CQ |
| Abdominal pain | 7 (11) | 3 (23) | 4 (9) | 0 (0) | 1 (11) | 2 (8) | 4 (13) |
| Anorexia | 2 (3) | 1 (8) | 1 (2) | 0 (0) | 1 (11) | 1 (4) | 0 (0) |
| Dizziness | 12 (18) | 2 (15) | 8 (19) | 2 (22) | 1 (11) | 3 (13) | 8 (25) |
| Fever | 10 (15) | 1 (8) | 7 (16) | 2 (22) | 1 (11) | 6 (25) | 3 (9) |
| Headache | 16 (25) | 5 (39) | 10 (23) | 1 (11) | 3 (33) | 6 (25) | 7 (22) |
| Nausea | 9 (14) | 1 (8) | 7 (16) | 1 (11) | 0 (0) | 4 (17) | 5 (16) |
| Palpitations | 1 (2) | 0 (0) | 1 (2) | 0 (0) | 0 (0) | 0 (0) | 1 (3) |
| Unknown | 4 (6) | 0 (0) | 3 (7) | 1 (11) | 1 (11) | 1 (4) | 2 (6) |
| Vomiting | 2 (3) | 0 (0) | 2 (5) | 0 (0) | 0 (0) | 1 (4) | 1 (3) |
| Weakness | 2 (3) | 0 (0) | 0 (0) | 2 (22) | 1 (11) | 0 (0) | 1 (3) |
| Total | 65 | 13 (20) | 43 (66) | 9 (14) | 9 (14) | 24 (37) | 32 (49) |
| Adverse events reported during three rounds of intermittent preventive treatment among primary school children in rural Blantyre, Malawi. IPT-DP, Intermittent Preventive Treatment with Dihydroartemisinin-Piperaquine; IPT-SP+CQ, Intermittent Preventive Treatment with Sulfadoxine-Pyrimethamine and Chloroquine. |  |  |  |  |  |  |  |

## Discussion

In this trial comparing three rounds of IPTsc-DP and IPTsc-SP+CQ to control (no IPTsc) during the peak malaria transmission period, both DP and SP+CQ protected school children from *Pf* infections, anemia, and clinical malaria episodes and increased hemoglobin concentrations. This is the first trial to evaluate the combination of SP and CQ for intermittent preventive treatment, showing that in school children, both DP and SP+CQ drug regimens reduce the burden of malaria. Due to the threat of artemisinin resistance and the increasing use of DP as a first-line treatment drug, the combination of SP+CQ could be an important regimen to consider for IPTsc.

As expected, we observed that IPTsc with DP significantly improved outcomes among participants compared to the control group. This is consistent with previous findings from a systematic review [13], as well as a more recent study in Tanzania [14] and the initial phase of this trial [29]. DP’s high efficacy is likely due to limited or no resistance to its component drugs and the long half-life of piperaquine, providing a prolonged period of prophylaxis against new infections [34]. DP is already widely used in Africa and comes in a fixed dose, co-packaged oral formulation. We observed fewer adverse events and better adherence compared to SP+CQ, suggesting DP may be more acceptable.

The combination of SP+CQ was also efficacious for IPTsc compared to control, though the combination was less effective than DP in reducing *Pf* infections. Based on the widespread re-emergence of chloroquine susceptible *Pf* parasites following withdraw of CQ from use and demonstrated return of clinical efficacy, we assume that CQ is highly effective with an estimated post-treatment duration of prophylaxis of ∼4 weeks [35–37]. Thus, the difference in efficacy between SP+CQ and DP in reducing *Pf* infections could be due to resistance to SP and/or shorter duration of prophylaxis compared to DP. Despite withdrawal of SP for treatment in Malawi, molecular markers of SP resistance remain prevalent [18, 26, 38], and the clinical utility of SP for chemoprevention is not clear. In pregnancy, IPT with SP remains effective at improving birth outcomes, though it is theorized that this is due to the antimicrobial or anti-inflammatory effects rather than the antimalarial efficacy [39]. Alternatively, SP may retain some antimalarial efficacy in semi-immune individuals without clinical infections [28, 40]. High concentrations of SP immediately after treatment, clear even highly SP-resistant parasites; however, the duration of prophylaxis may be compromised by both recrudescent parasites as well as reinfection with resistant parasites [37]. Quantifying the parasitological efficacy and duration of post-treatment prophylaxis of SP for chemoprevention in settings like Malawi is critical for informing drug choice for both IPTsc and chemoprevention targeting younger children, i.e. seasonal or perennial malaria chemoprevention. If SP has limited or no parasitological efficacy, then partner drugs (e.g. CQ or amodiaquine) are being used as monotherapy [41]. Thus, while our results are promising, more consideration is required before recommending SP+CQ for widespread use.

Overall, both IPTsc-DP and IPTsc-SP+CQ were safe and well tolerated, as there were no serious adverse events (AEs). Similar to results from the initial study, most AEs, which were grade 1 and 2, occurred in participants who received CQ. Hydroxychloroquine, the less toxic metabolite of CQ, was used in this study. The well-established safety profile for CQ is a significant advantage for use in girls of childbearing potential, who would often be left unprotected by other regimens, but could be a particularly important target population. Clearing and preventing infections in adolescent girls may improve pre-conception health of high-risk adolescent pregnancies and prevent first-trimester malaria [42].

While this trial was successfully implemented and had good participant follow-up, some limitations may affect the interpretation and generalizability of our findings. First, attrition between the first and second phases of the study may have partially compromised the original randomization and introduced post-randomization selection (attrition) bias; however, most losses were due to school completion rather than loss to follow-up and were therefore likely unrelated to the treatment assignment or study outcomes. Second, children allocated to the IPTsc-DP and control arms had the same treatment experience in both trial phases, while children in the IPTsc-SP+CQ arm had a different treatment experience in the initial study. However, at baseline for the continuation study, the three study groups were comparable, reducing the likelihood that the differing treatment effects in the initial study impacted our findings. Third, the lack of a placebo control group could have led to underestimation of the effect of IPTsc-DP and IPTsc-SP+CQ on clinical malaria due to altered treatment-seeking behavior in the control arm from a recognition of not getting any treatment. However, all participants were instructed to seek treatment at the health center if they were symptomatic. In addition, there was potential for performance bias in outcome assessments due to the lack of blinding in participants and assessors. However, the primary outcome assessments were done by laboratory technicians who were blinded to the study allocation. Another consideration is that 82% of our participants got three doses of treatment, and this can be challenging in a real-world setting with less capacity for participant follow-up, which risks reduced coverage and lower impact of IPTsc on infection and clinical malaria episodes. A single-dose drug for chemoprevention could dramatically improve compliance and feasibility. Finally, the results of this study are specific to the drug resistance and transmission patterns in Malawi and similar settings, thus may not be generalizable to places with different drug resistance profiles and malaria transmission patterns.

In summary, our study adds to the growing body of evidence supporting the World Health Organization’s recommendation for IPTsc in areas of moderate to high malaria transmission. Importantly, our study demonstrates that SP+CQ, an alternative non-artemisinin combination regimen, can be effective for IPTsc. Further research either in trials or implementation settings is needed to understand the broader impact of IPTsc on malaria transmission and education. In settings like Malawi where DP may not be suitable for IPTsc due to its role as an alternative first-line treatment for uncomplicated malaria, SP+CQ may be a viable and practical alternative.

## Data Availability

Data are in the process of submission to the Infectious Disease Data Observatory (IDDO - https://www.iddo.org/wwarn/how-share-data)

## Acknowledgements

We would like to thank the students of Nainunje Primary School and their guardians/parents for participating in the study. We also thank the staff of Nainunje Primary School and the surrounding community for allowing the study to be conducted in this area. We thank Dr. Charles Mangani who served as our local safety monitor. Finally, we would like to thank the Malaria Alert Centre study team who recruited participants, collected data, samples and conducted participant follow-up.

## Notes

### Competing Interest Statement

The authors have declared no competing interest.

### Clinical Trial

ClinicalTrials.gov ID: NCT05980156 https://clinicaltrials.gov/study/NCT05980156

### Author Declarations

The study protocol was reviewed and approved by the Kamuzu University of Health Sciences Research Ethics Committee (P.06/21/3410) and the University of Maryland School of Medicine (HCR-HP-00098250-2)

